# Multiplex RT-qPCR assay (N200) to detect and estimate prevalence of multiple SARS-CoV-2 Variants of Concern in wastewater

**DOI:** 10.1101/2022.04.12.22273761

**Authors:** Meghan Fuzzen, Nathanael B.J. Harper, Hadi A. Dhiyebi, Nivetha Srikanthan, Samina Hayat, Shelley W. Peterson, Ivy Yang, J.X. Sun, Elizabeth A. Edwards, John P. Giesy, Chand S. Mangat, Tyson E. Graber, Robert Delatolla, Mark R. Servos

**Affiliations:** Department of Biology, University of Waterloo, Waterloo, ON, N2L 3G1, Canada; One-Health Division, Wastewater Surveillance Unit, National Microbiology Laboratory, Public Health Agency of Canada, Winnipeg, MB, R3E 3M4, Canada; Chemical Engineering and Applied Chemistry, University of Toronto, Toronto, ON, M5S 3E5, Canada; Department of Veterinary Biomedical Sciences and Toxicology Centre, University of Saskatchewan, Saskatoon, SK S7N 5B3, Canada; Department of Environmental Sciences, Baylor University, Waco, TX, USA; Department of Zoology and Center for Integrative Toxicology, Michigan State University, East Lansing, MI, USA; Children’s Hospital of Eastern Ontario Research Institute, Ottawa, Ontario, K1H 8L1, Canada; Department of Civil Engineering, University of Ottawa, Ottawa, Ontario, K1N 6N5, Canada

## Abstract

Wastewater-based surveillance (WBS) has become an effective tool around the globe for indirect monitoring of COVID-19 in communities. Quantities of viral fragments of SARS-CoV-2 in wastewater are related to numbers of clinical cases of COVID-19 reported within the corresponding sewershed. Variants of Concern (VOCs) have been detected in wastewater by use of reverse transcription quantitative polymerase chain reaction (RT-qPCR) or sequencing. A multiplex RT-qPCR assay to detect and estimate the prevalence of multiple VOCs, including Omicron/Alpha, Beta, Gamma, and Delta, in wastewater RNA extracts was developed and validated. The probe-based multiplex assay, named “N200” focuses on amino acids 199-202, a region of the N gene that contains several mutations that are associated with variants of SARS-CoV-2 within a single amplicon. Each of the probes in the N200 assay are specific to the targeted mutations and worked equally well in single- and multi-plex modes. To estimate prevalence of each VOC, the abundance of the targeted mutation was compared with a non-mutated region within the same amplified region. The N200 assay was applied to monitor frequencies of VOCs in wastewater extracts from six sewersheds in Ontario, Canada collected between December 1, 2021, and January 4, 2022. Using the N200 assay, the replacement of the Delta variant along with the introduction and rapid dominance of the Omicron variant were monitored in near real-time, as they occurred nearly simultaneously at all six locations. The N200 assay is robust and efficient for wastewater surveillance can be adopted into VOC monitoring programs or replace more laborious assays currently being used to monitor SARS-CoV-2 and its VOCs.

## Introduction

Communities around the world have quickly adopted wastewater-based surveillance (WBS) of SARS-CoV-2 viral fragments to detect and infer prevalence of COVID-19 disease burden in communities (D’Aoust et al., 2021; Kitajima et al., 2020; Medema et al., 2020). Because it does not require testing of individual sensitivity and includes all individuals, symptomatic or asymptomatic, that contribute to the sewershed, WBS has several advantages over more costly and invasive clinical testing. WBS utilizes reverse transcription quantitative real-time polymerase chain reaction (RT-qPCR) based techniques targeting various regions of the SARS-CoV-2 genome, including the popular US-Centers for Disease Control (CDC) N1, N2 (nucleocapsid) and E (envelope), to test for the presence of SARS-CoV-2 in wastewater (Hamouda et al., 2021; Kitajima et al., 2020; Kumblathan et al., 2021).

Continuous emergence of Variants of Concern (VOCs) and Variants of Interest (VOIs) has presented additional challenges to the surveillance of SARS-CoV-2 and has been an area where WBS can provide more timely information quicker than clinical testing. In addition to clinical tracking of abundance of SARS-CoV-2 infections, RT-qPCR methods have been used to detect and monitor prevalence of emerging VOCs in wastewater (Carcereny et al., 2021; Graber et al., 2021; Lee et al., 2021; Peterson et al., 2022; Wurtzer et al., 2021). This information provides an integrated estimation of the relative abundance of variants within populations. There have been several examples of RT-qPCR assays for identification and quantifications of VOCs wastewater. The mutations N501Y and Δ69-70 del in the spike protein or the presence of mutation D3L in the N gene have been targeted by RT-qPCR assays to monitor for the presence of VOCs in wastewaters across Canada (Peterson et al., 2022). Using an allele specific RT-qPCR, with an artificial mismatch, three mutations, HV69/70del, Y144del and A570D, were targeted to discriminate and quantify the Alpha from VOC relative to the wild type in wastewater of 19 communities in the USA (Lee et al, 2021). Another assay targets mutations Δ69-70 in the S gene to detect the Alpha and Beta variants respectively (Yaniv et al., 2021). An assay based on mutation D3L of the gene, was applied for detection and quantification of relative proportion of the Alpha VOC (Graber et al., 2021). Those authors were able to effectively trace the increase of the proportion of Alpha variant in wastewater of Ottawa, Canada, in almost real time. The trends in incidence of Alpha were similar to that of the number of clinical cases in the community. These various studies demonstrate that WBS can rapidly provide information on incidence and prevalence of SARS-CoV-2 to authorities with sensitivity and lineage specificity. RT-qPCR assays are specific and can be deployed rapidly across existing WBS networks. The challenges with using RT-qPCR assays in surveillance of VOCs are the amount of time needed to develop and validate new assays each time a new VOC emerges as well as increasing the number assays being conducted to continually monitor for various VOCs and VOIs.

The objective of this study was to develop a multiplex RT-qPCR assay that could be used to simultaneously screen for several VOCs, as well as a marker of the total SARS-CoV-2 present in the sample so that an estimation of the relative proportion of VOCs can be reliably determined. The N gene region was selected for several reasons, including that this region is targeted by the N1 and N2 assays, developed by the US CDC, which are commonly used for monitoring wastewater (Lu et al., 2020). There is evidence to suggest that assays developed for the N-region of the SARS-CoV-2 genome have greater sensitivity for measuring various VOCs of SARS-CoV-2 in wastewater than assays targeting than does the S-(Yaniv et al., 2021) or the envelope regions (Pérez-Cataluña et al., 2021). The N-gene was selected as the target region as the target of the assay since it is also rich in mutations unique to VOCs (Kiryanov et al., 2022). Specifically, the 121-basepair region in the N-gene open reading frame (ORF) (nucleotides 28,837 to 28,958) that includes single or multiple nucleotide variants for each VOC was selected (Table 1). According to GISAID data summarized by nextrain.org (Hadfield et al., 2018), the nucleotide 28880- 28882 (AA N:203) have one of the highest entropy levels in the SARS-CoV-2 genome, at 0.972, with nucleotides 28883-28885 (AA N:204) also having high entropy level of 0.711 (nextstrain.org; accessed April 1, 2022). The non-synonymous functional polymorphisms present in this region include R203M, present in Delta, which increases the spread of the virus (Syed et al., 2021); R203K and G204R, (both present in Alpha, Gamma Lambda, and Omicron, which are related to increased infectivity, fitness and virulence (Johnson et al., 2021; Lee et al., 2021; Wu et al., 2021); and T205I, present in Beta and Mu variants that were predicted to have reduced antigenicity and greater affinity for HLA-I (Antonio et al., 2021). The presence of multiple mutations in a single region allowed for the design of a single amplicon, multiple probe assay, that can distinguish among several VOCs. A Universal probe, that targets an area of the amplicon without mutations allows for the quantification of the total SARS-CoV-2 signal, regardless of which variants are present in wastewater. This allows for the measurement of the total SARS-CoV-2 signal as well as providing estimations of the relative abundance of individual VOCs.

**Table 1.** Amino acid mutations and corresponding nucleotide mutations present in the targeted region of the SARS-CoV-2 nucleocapisd genome (AA 199-214).

| Amino acid mutation(s) | Nucleotide mutation(s) | Variant(s) of concern with mutation(s) | Mutation Prevalence (# of GISAID sequences) |
| --- | --- | --- | --- |
| S202S<br>(synonymous mutation) | A28877T<br>G28878C | Gamma | 98.0% and 97% respectively<br>(6,925,348) |
| R203K | G28881A<br>G28882A | Alpha<br>Gamma<br>Omicron | 97.8% (1,144,688)<br>94.5% (120,031)<br>98.5% (295,436) |
| R203M | A28880T | Delta | 98.7% (3,984,837) |
| G204R | G28883C | Alpha<br>Gamma<br>Omicron | 90.8% (1,144,688)<br>94.9% (120,031)<br>98.6% (295,496) |
| T205I | C28886T | Beta | 96% (40,518) |

<sup>A</sup> Mutation prevalence was obtained from outbreak.info on January 14, 2022.

Prior to being incorporated into a wastewater surveillance program, the N200 assay was validated using several synthetic standards as well as samples of wastewaters. As an example of the utility of this assay, between November and January 2021, samples from six wastewater treatment facilities in Ontario, Canada, were examined with the N200 assay. During this period the N200 assay was utilized to test for the presence and increased frequency of the R203K/G204R mutation (present in the Omicron VOC), while comparing directly to the decrease in frequency of the R203M mutation (present in the Delta VOC), relative to a VOC-agnostic Universal marker, all within a single RT-qPCR reaction.

## Materials and Methods

### Target selection

To select an appropriate RT-qPCR target for testing wastewater, mutations that were indicative of each VOC, encompassing amino acids N:202-205 were identified. Mutations selected for each VOC and their known prevalence are presented in Table 1. The template sequence used for designs were retrieved from accession numbers provided by TWIST Bioscience (South San Francisco, CA, USA) for synthetic controls 14 (Alpha), 16 (Beta), 17 (Gamma) and 23 (Delta) and sequences were aligned using MAFFT (Katoh et al., 2002).

### Assay design

Candidate forward and reverse primers that encompassed an amplicon 121 base pairs long containing all the desired mutations were designed with a target predicted T_m_ around 61°C and 40-60% GC content. Candidate primers were screened for hairpins, homodimers and heterodimers *in silico* using OligoAnalyzer by integrated DNA technologies (IDT; RRID:SCR_001363) with the “qPCR” parameter selected. Candidate primers that passed were then screened against NCBI’s non-redundant nucleotide database using BLAST (Altschul et al., 1990) and Primer-BLAST (Ye et al., 2012) to maximize mismatches with as much non-target genetic material as possible (human, microbes, food, etc.).

Five probes were designed to be selective for each of the mutations (Table 1, 2), as well as a universal probe for the “Universal” probe complementary to a nearby, highly conserved (bp 28891-28905, mutation rates <0.87% according to covidcg.org) region of the amplicon to detect total SARS-CoV-2 (Table 2, Figure 1). Probes were designed with the mutated base(s) at their center using PrimerExpress v3.0.1 with a GC content of 40-60%. BHQplus technology (LCG, Biosearch Technologies, Inc., California, USA) was used to increase the T_m_ of each probe, to have a predicted T_m_ of 69°C, which enabled the design of short probes approximately 15 nucleotides long. Probes were then tested for hairpins, homodimers, and heterodimers. Candidate probes were also screened for % identity to non-target sequences using NCBI’s BLAST tool (Agarwala et al., 2016).

**Table 2.**
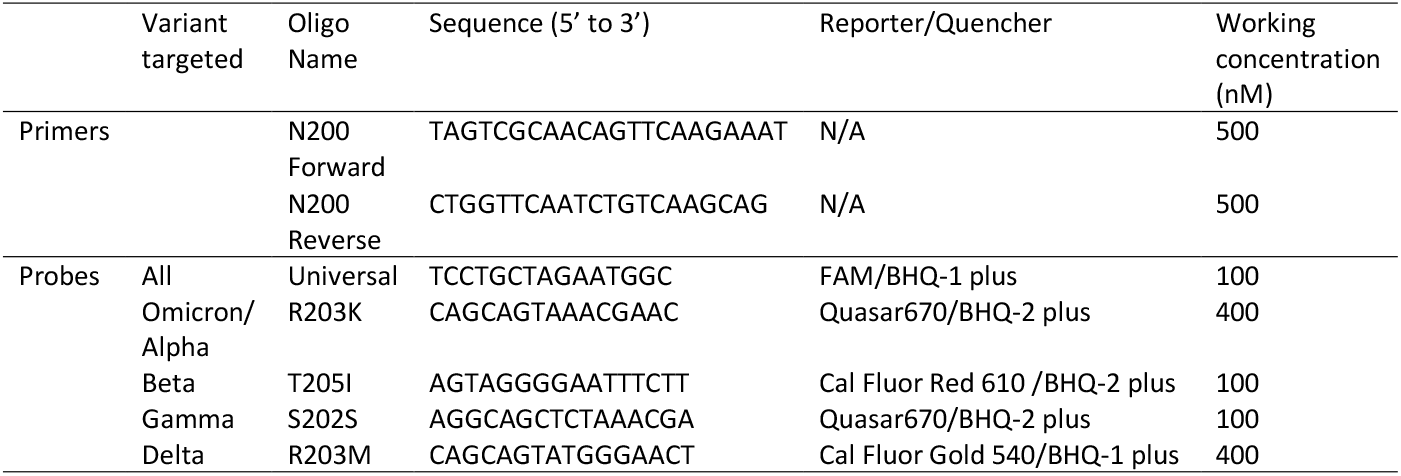
Forward (F) and Reverse (R) primer and probe designs for the N200 VOC assay. The reporter and quencher moieties, and final working concentrations are included for each oligonucleotide.

**Figure 1.**
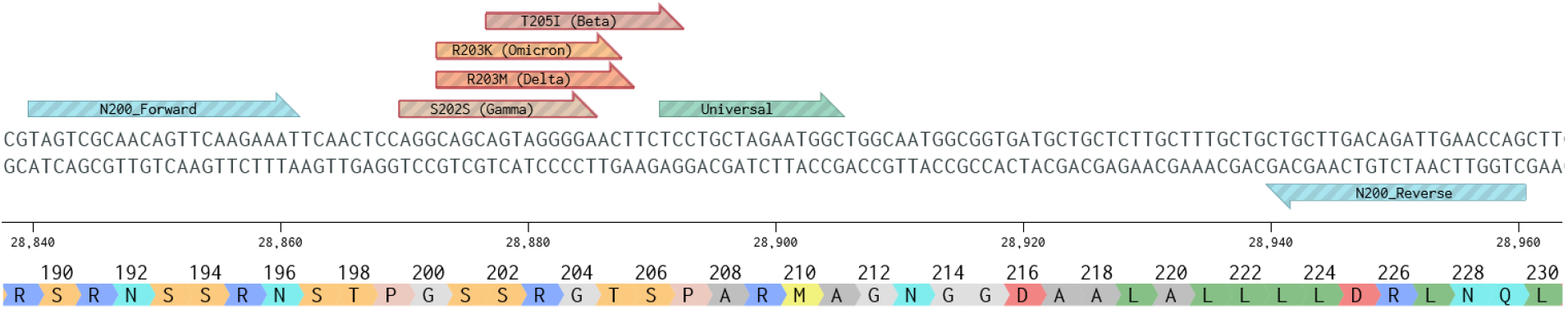
N200 assay amplicon with the location of the N200 forward and reverse primers and all probes are shown on a portion of the N gene of the SARS-CoV-2 genome. The sequence, nucleotide numbers, and ammino acid displayed are based on the SARS-CoV-2 reference genome (Wuhan; NC_045512.2; figure generated using Benchling.com).

### Assay Validation

Initial tests of the N200 assay were performed on synthetic templates. TWIST synthetic RNA controls 2, 14, 16, 17, and 23 (TWIST Bioscience) were employed to generate standard curves (sequences representative in lieu of Wuhan-Hu-1 (Genbank MN908957.3), as well as the VOCs, Alpha, Beta, Gamma, and Delta VOCs, respectively). The N200 standard curves was assessed for linearity and efficiency by use of each of the probes in singleplex. After validation with synthetic templates assays were tested with wastewater RNA extracts to ensure successful amplification. Temperature gradients and primer/probe concentration gradients were performed to determine the optimal assay conditions in singleplex.

Cross-reactivity of probes was assessed by use of synthetic RNA controls. Each probe, in singleplex, was tested for reactivity with TWIST RNA controls 2, 14, 16, 17 and 23 at a concentration of 500 copies per reaction. Specificity was also determined in a triplex assay iteration (Universal, R203M, and R203K probes) by use of digital PCR (dPCR) at multiple concentrations of TWIST RNA controls (see dPCR section for additional details).

The N200 assay was validated in multiplex by comparing the efficiency, R^2^ and y- intercept of standard curves, as well as the estimated concentration and proportion of variants tested in wastewater relative to singleplex. Sensitivity of the N200 assay with the Universal, R203K, and R203M probes were determined by use of a 12-point standard curve (TWIST synthetic controls) with 15-replicates each. The limit of detection (LOD) was defined as the standard concentration at which >95% of the replicates were detected. This was performed by two independent labs (at the University of Waterloo and the Canadian National Microbiology Laboratory – Winnipeg) using the same suppliers of primers and probes and standards.

### Wastewater sample collection, concentration, and extraction

Wastewater from the Regions of Peel (wastewater treatment plant influents GE Booth and Clarkson), York (access points Humber Air Management Facility (AMF) and Warden) and Waterloo (wastewater treatment plant influents Kitchener and Waterloo) Ontario, Canada. At each site wastewater was collected by plant operators using 24-hour cooled composite samplers (combined sample of three grabs at Warden) 3-5 times a week between November 28^th^, 2021, and January 4^th^, 2022. Samples were aliquoted into 250 mL HPDE bottles (Systems Plus, Baden, Canada) and transported on ice to the University of Waterloo.

To concentrate RNA, 250 mL samples were well-mixed by inversion, 40 mL aliquots poured into 50 mL conical tubes with 4 g PEG-8000 and 0.9 g NaCl, then mixed on an orbital shaker at low speed at 4°C for 2 h before being stored at 4°C overnight (Wu et al., 2020). Samples were then centrifuged at 12,000 x g at 4°C for 90 minutes with no brake. The supernatant was discarded, and the remaining sample was centrifuged again at 12,000 x g at 4°C for 5 min with no brake. The remaining supernatant was decanted and pipetted out and the wet weight of the resulting pellet recorded. Up to 250 mg of the pellet was used for extraction of RNA by use of the RNeasy PowerMicrobiome Kit (Qiagen, Germantown, MD) as per manufacturers instructions and included the addition of 100 µL of TRIzol (Thermo Fisher, Mississauga, Canada) to the pellet before bead-beating. Total RNA was eluted in 100 µL of RNase-free water.

### RT-qPCR

One-step RT-qPCR was performed using TaqPath 1-Step Master Mix (Thermo Fisher, Mississauga, Canada). The primer and probe sequences and final working concentrations are presented in Table 2. RT-qPCR reactions were run in triplicate using 5 µL RNA template and a final reaction volume of 20 µL. Cycling was performed on an OPUS or CFX 96 Touch qPCR thermocycler (BioRad, Hercules, CA) as follows: RT at 50°C for 15 min, 95°C for 2 min, 45 cycles of 95°C for 3 sec followed by 57°C for 30 sec. All qPCR plates were run with controls including no template controls (NTCs), a positive control (wastewater sample with previously determined amount of Delta variant) and a standard curve based on a homologous synthetic RNA (i.e., EDX, TWIST).

### Digital PCR (dPCR)

For precise quantification, commercial standards were analyzed via singleplex one-step RT-dPCR (QIAcuity, Qiagen, Hilden, Germany). QIAquity One-Step Viral RT-PCR Kit (Qiagen) together with CDC_N1 probe and primers (Lu et al., 2020) were used to determine absolute copy number (cp) of RNA in the following SARS-CoV-2 genomic RNA standards: TWIST Bioscience Controls 2, 14, 16, 17, 23. Testing for specificity of probes was also conducted using the N200 assay in triplex (Universal, R203M, and R203K probes) with two concentrations of TWIST Bioscience Controls 14, and 23. Reactions consisted of 10 µL of RNA template, forward and reverse primers (working concentration of 500 nM each), probe (working concentration of 125 nM for N1, or 100, 400 and 400 nM for the Universal, R203M and R203K probes respectively), 10 µL of supermix, 0.4 µL of reverse transcriptase, and 14.6 µL of PCR-grade water to make up the balance of the 40 µL reaction. All reactions were performed in triplicate using 24 well x 26,000 channel plates. Cycling was performed on the QIAcuity as follows: RT at 50°C for 30 min, 95°C for 2 min, with 40 cycles at 95°C for 3 sec then annealing at 55°C or 57°C (for N1 and N200 assays respectively) for 30 sec. The N200 assay was also tested on RT-dPCR in triplex (Universal, R203M, and R203K probes) using the TWIST Bioscience Controls 23 and 48 at multiple concentrations to determine the specificity of the assay.

### Data Analysis

Samples were quantified (copies/well) by use of a 7-point standard curve with the relevant TWIST standard. Samples were then corrected for elution volume and volume of wastewater extracted and reported as copies/mL wastewater (Quantity). The proportion of total Quantity associated with each VOC in a sample was determined by use of the equation mutation specific for the VOC (Equation 1).

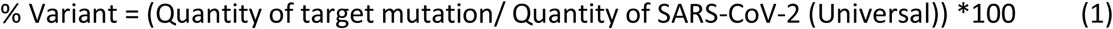

## Results

### N200 assay conditions and performance

The N200 assay performed well with all probes in singleplex. When tested with a serial dilution of TWIST standards, the singleplex standard curves were linear, had an efficiency near 100%, and the slopes and y-intercepts were comparable between each of the probes (Table 3). The N200 assay performed similarly when tested in singleplex, duplex and triplex (Table 3). All probes targeting mutations were found to be highly specific with little to no cross-reactivity for non-target templates using qPCR (in singleplex; Table 4). Using RT-qPCR, this was demonstrated by the lack of amplification on all VOC TWIST templates without the target mutation (Table 4). Some cross-reactivity was observed between the T205I probe and the Wuhan variant control (TWIST Control 2; Table 4), but this accounted for less than 5% of the signal observed on the homologous template. Positive amplification with similar sensitivity was seen on of all TWIST templates tested was observed with the Universal probe (Table 4). High target specificity of each probe was confirmed by RT-dPCR, where in the presence of a high concentration of non-target template, almost no amplification was detected (Table 5). Copy numbers reported by the mutation probes (R203M, R203K) were comparable to the copy numbers reported by the Universal probe in the RT-dPCR reactions at multiple concentrations, which further suggests that each probe has similar sensitivities and efficiencies (Table 5). The LOD (95% detection) was determined to be 4-6 copies per reaction for the Universal, R203K, and R203M probes respectively when tested in two separate laboratories.

**Table 3.** Performance metrics of the N200 VOC assay for each of the probes in singleplex, duplex or triplex.

|  | Assay | Slope | y-intercept | Efficiency | R<sup>2</sup> |
| --- | --- | --- | --- | --- | --- |
| Universal | Singleplex | -3.347 | 40.236 | 98.9 | 0.990 |
|  | Duplex (with R203M) | -3.366 | 39.50 | 98.2 | 0.991 |
|  | Duplex (with R203K) | -3.299 | 38.793 | 101.0 | 0.990 |
|  | Duplex (with S202S) | -3.311 | 40.207 | 100.5 | 0.990 |
|  | Duplex (with T205I) | -3.461 | 39.773 | 94.5 | 0.990 |
|  | Triplex (with R203M, R203K) | -3.443 | 39.661 | 95.2 | 0.995 |
| R203K<br>(Omicron/Alpha) | Singleplex | -3.429 | 38.834 | 95.7 | 0.998 |
|  | Duplex (with Universal) | -3.334 | 39.585 | 99.5 | 0.991 |
|  | Triplex (with Universal, R203M) | -3.464 | 39.902 | 94.4 | 0.995 |
| T205I<br>(Beta) | Singleplex | -3.396 | 39.951 | 97.0 | 0.997 |
|  | Duplex (with Universal) | -3.533 | 39.952 | 91.9 | 0.994 |
| S202S<br>(Gamma) | Singleplex | -3.249 | 40.363 | 103.1 | 0.998 |
|  | Duplex (with Universal) | -3.352 | 39.891 | 98.8 | 0.988 |
| R203M<br>(Delta) | Singleplex | -3.541 | 40.365 | 91.6 | 0.998 |
|  | Duplex (with Universal) | -3.387 | 39.513 | 97.4 | 0.990 |
|  | Triplex (with Universal, R203K) | -3.442 | 39.124 | 95.2 | 0.995 |

**Table 4.**
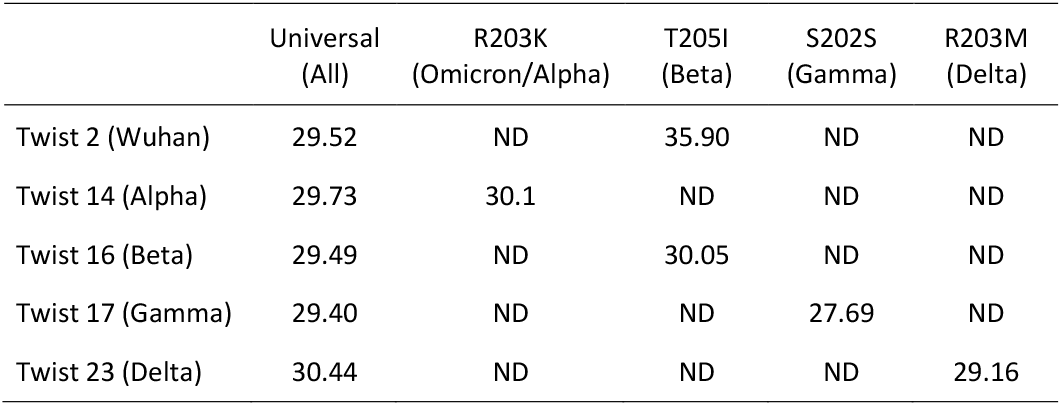
Cycle Threshold (CT) value (mean) of N200 assay performed in singleplex with probes targeting a mutation (R203K; T205I; S202S; R203M) or a common region (Universal) of the SARS-CoV-2 genome. Individual probes were performed on each of five synthetic templates synthesized by TWIST Bioscience (San Francisco, USA), controls for the original Wuhan strain and for each VOC. ND = not detected

|  | Universal<br>(All) | R203K<br>(Omicron/Alpha) | T205I<br>(Beta) | S202S<br>(Gamma) | R203M<br>(Delta) |
| --- | --- | --- | --- | --- | --- |
| Twist 2 (Wuhan) | 29.52 | ND | 35.90 | ND | ND |
| Twist 14 (Alpha) | 29.73 | 30.1 | ND | ND | ND |
| Twist 16 (Beta) | 29.49 | ND | 30.05 | ND | ND |
| Twist 17 (Gamma) | 29.40 | ND | ND | 27.69 | ND |
| Twist 23 (Delta) | 30.44 | ND | ND | ND | 29.16 |

**Table 5.** Gene copies/µL of RNA (mean ± SD) detected by the Universal, R203M, or R203K probes in the N200 assay (triplexed) when tested against TWIST controls for variants of SARS-CoV-2 using RT-dPCR.

| | Dilution from stock<br>($\sim 1 \times 10^4$ copies/$\mu\text{L}$) | Universal<br>(All) | R203K<br>(Omicron/Alpha) | R203M<br>(Delta) |
| --- | --- | --- | --- | --- |
| Twist 23<br>(Delta) | 5x | 2769 ± 173 | 0 ± 0 | 2767 ± 173 |
| | 500x | 25 ± 2 | 0 ± 0 | 25 ± 2 |
| | 5000x | 3 ± 1 | 0 ± 0 | 3 ± 1 |
| Twist 48<br>(Omicron) | 5x | 824 ± 12 | 820 ± 22 | 2 ± 2 |
| | 50x | 78 ± 3 | 77 ± 3 | 0 ± 0 |
| | 500x | 5 ± 0 | 5 ± 0 | 0 ± 0 |

### Performance of N200 assay with samples of wastewater

The triplex N200 assay was applied to wastewater samples throughout December 2021 and January 2022 to monitor for community transmission of the Omicron VOC, which would be signified by decrease in the proportion of the R203M (Delta) mutation and an increase in the R203K mutation (Omicron). The total SARS-CoV-2 (Universal) signal in samples during this period started out low, but rapidly increased over the sampling period (Figure 2). During this time, we observed a rapid transition in relative abundance of mutations from being dominated by R203M (Delta) to being dominated by R203K (Omicron) at all monitoring locations (Figure 3).

**Figure 2.**
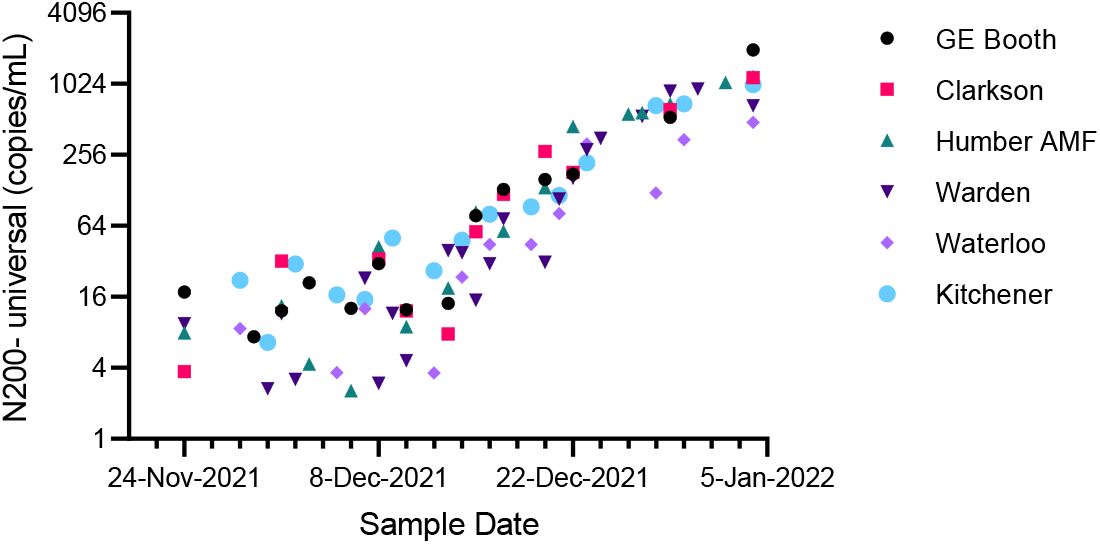
SARS-CoV-2 signal measured in municipal wastewater using the N200 assay with the Universal probe in a multiplexed rection (Log 2 scale – copies/mL). Samples from the Peel (GE Booth and Clarkson), York (Humber AMF and Warden) and Waterloo (Waterloo and Kitchener) Regions were collected during the period encompassing both the introduction of Omicron into these communities and the concomitant reduction of the endemic Delta VOC.

**Figure 3.**
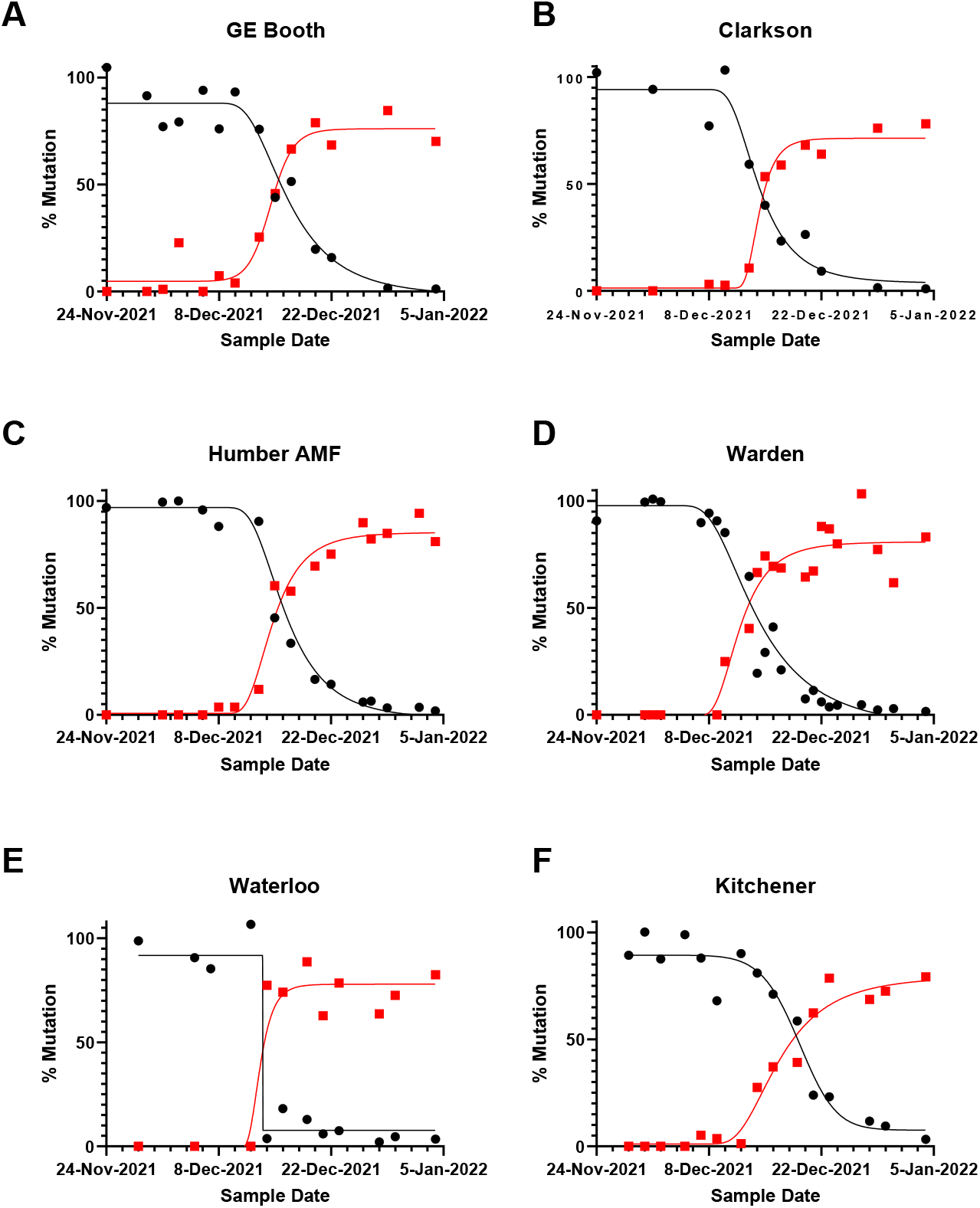
Application of the N200 triplex assay to monitor the relative abundance (%) of the mutations R203M (black circles- presumed Delta) or R203K/G204R (red squares – presumed Omicron) in municipal wastewater effluent. Effluent was collected from the Region of Peel (A, B), York Region (C, D) and Waterloo Region (E, F) using composite (A-B, D-F) and grab (C) sampling. Lines drawn through points are a 5-point sigmoidal curve calculated using GraphPad PRISM (v 9.3.1).

## Discussion

This study presents an RT-qPCR approach for detection of SARS-CoV-2 and VOCs Omicron/Alpha, Beta, Gamma and Delta in a multiplexed assay with a single amplicon. The specificity and sensitivity of the N200 assay allow for simultaneous detection and quantification of mutations associated with VOCs. These individual assays were multiplexed and validated in duplex and triplex reactions, which allowed for rapid, cost-effective detection and quantitation of mutations associated with VOCs. Relative amounts of these mutations are then compared to a non-mutated region of the same amplicon to estimate the prevalence of current VOCs in a wastewater sample. This assay was able to track the initial presence and then the transition from the R203M mutation (i.e., Delta) as it was replaced by the R203K/G204R mutation (i.e., Omicron/Alpha) in multiple communities in south-central Ontario in December 2021. This is representative of the transition from the Delta variant to the Omicron variant observed in clinical cases at this time in these communities.

In addition to R203M (Delta) and R203K/G204R (Omicron/Alpha) targets, probes targeting the T205I (Beta) mutation and the silent mutation in the amino acid S202 (Gamma), allow for surveillance of known VOCs using a single assay. Having multiplexed, variant specific probes that are compared to a Universal reference within the same amplicon is beneficial for improved consistency and accuracy of measuring relative prevalence of variants. As the gene targets are all in the same 121 bp amplicon, there is minimal concern about the dynamics of different regions of the SARS-CoV-2 genome in either the wastewater system itself, or in the extraction process. Because the reactions occur together in a multiplexed assay there is reduced variability and replication compared to running assays separately. This approach saves resources and time, as fewer reactions are run per sample. Critically, this assay also saves sample volume, which is often limited, allowing for additional assays to be run on the same extract or reducing the need for multiple extractions to be performed. Ultimately it makes the process more efficient, reduces turnaround time and improves quality.

The combined use of Universal and VOC-specific probes in the N200 assay to estimate prevalence of VOCs in wastewater is advantageous for several reasons. First, since when multiplexed, the mutation and comparator are being assessed within the same amplicon as well the same reaction, this method provides increased precision of VOCs prevalence estimates. Additional confidence in prevalence of VOCs is provided by the fact that the mutation and Universal signals can be quantified using the same standard. In comparison, other VOC qPCR assays estimate prevalence by comparing a mutant allele to a genetic marker of SARS-COV-2 in another amplicon, /genetic locus such as the CDC_N1 (Vogels et al., 2021; Wolfe et al., 2021), or by comparing the frequencies of the mutant and “wild-type” (e.g., ancestral/endemic allele) at the same locus (Chan et al., 2022; Graber et al., 2021; Lee et al., 2021; Peterson et al., 2022). While these assays can produce accurate data (from both analytical and clinical perspectives), there are additional controls needed to account for variations in efficiencies of variation (inter-assay variability and matching standard quantities), that are eliminated by the N200 assay. Estimations of VOC prevalence are important during wastewater surveillance SARS-CoV-2 in that they can provide useful information to public health units in a timely manner.

During analyses of wastewaters through late 2021 and early 2022, a rapid increase in the SARS-CoV-2 signal (Universal probe) as well as a quick change over of the dominant variant from Delta (R203M) to Omicron (R203K/G204R) was observed at all sites monitored. The assay first detected the R203K mutation in a sample collected from GE Booth (Peel Region) on December 3^rd^, 2021. While it is possible that this early detection of the R203K/G204R mutation could have been a variant other than Omicron, within a week of this detection, the R203K/G204R mutation was consistently detected in the wastewater at this site while R203M decreased in proportion. The use of two mutation targets provided confidence in the data as it was being collected and reported because both a decrease in the proportion of the R203M mutation (Delta) and an increase in the proportion of the R203K/G204R mutations (Omicron) occurred simultaneously. This data was reported to public health partners within a week of samples being collected and this was immediately disseminated to the public by the Public Health Unit as part of a weekly Epi-Report (https://data.peelregion.ca/documents/profile-of- covid-19-cases/explore). Rapid emergence of Omicron presented a major challenge for clinical testing and sequencing capacity. This was also confounded by the holiday season in Ontario. The wastewater data in general, quickly became important to Public Health Units as the clinical testing became less reliable because of changes to testing eligibility in Ontario. They were able to confirm the rapid replacement of Delta with the Omicron variant using wastewater that was independent of clinical testing (Arts et al., 2022).

The N200 multiplex qPCR was developed to improve estimates of the proportion of various variants in wastewater, while also making the assay more efficient to use in terms of reduced use of extracts, reagents, and time. The assay was validated using synthetic templates and was successfully applied to monitor for the emergence of the Omicron variant in several communities in Ontario. Although, this assay is one among several now available for detection of VOCs in wastewater, the unique design, with a single amplicon, multiple target mutations, and universal comparator targeting a highly mutable area of the N-gene make it very useful in wastewater-based surveillance of SARS-CoV-2.

## Data Availability

All data produced in the present work are available upon reasonable request to the authors.

## Acknowledgements

Funding for this project was provided by the Ontario Ministry of the Environment, Conservation and Parks, and NSERC Discovery and Canada Research Chairs. The research published in this paper is part of the project titled “Next generation solutions to ensure healthy water resources for future generations” funded by the Global Water Futures program, Canada First Research Excellence Fund. Additional information is available at www.globalwaterfutures.ca. The Municipal and Public Health Unit staff contributed to sample collection and provided valuable advice. Codey Dueck (PHAC) for laboratory analysis supporting LOD determinations. Yash Badlani, Carly Sing-Judge, Emily Dodsworth and Samantha Maerten assisted with sample preparation and analysis.

## References

Agarwala, R., Barrett, T., Beck, J., Benson, D.A., Bollin, C., Bolton, E., Bourexis, D., Brister, J.R., Bryant, S.H., Canese, K., Charowhas, C., Clark, K., Dicuccio, M., Dondoshansky, I., Federhen, S., Feolo, M., Funk, K., Geer, L.Y., Gorelenkov, V., Hoeppner, M., Holmes, B., Johnson, M., Khotomlianski, V., Kimchi, A., Kimelman, M., Kitts, P., Klimke, W., Krasnov, S., Kuznetsov, A., Landrum, M.J., Landsman, D., Lee, J.M., Lipman, D.J., Lu, Z., Madden, T.L., Madej, T., Marchler-Bauer, A., Karsch-Mizrachi, I., Murphy, T., Orris, R., Ostell, J., O’sullivan, C., Panchenko, A., Phan, L., Preuss, D., Pruitt, K.D., Rodarmer, K., Rubinstein, W., Sayers, E., Schneider, V., Schuler, G.D., Sherry, S.T., Sirotkin, K., Siyan, K., Slotta, D., Soboleva, A., Soussov, V., Starchenko, G., Tatusova, T.A., Todorov, K., Trawick, B.W., Vakatov, D., Wang, Y., Ward, M., Wilbur, W.J., Yaschenko, E., Zbicz, K., 2016. Database resources of the National Center for Biotechnology Information. Nucleic Acids Res. 44, D7. https://doi.org/10.1093/NAR/GKV1290

Altschul, S.F., Gish, W., Miller, W., Myers, E.W., Lipman, D.J., 1990. Basic local alignment search tool. J. Mol. Biol. 215, 403–410. https://doi.org/10.1016/S0022-2836(05)80360-2

Antonio, M., Pretti, M, Galvani, R.G., Farias, A.S., Boroni, M., Antônio, M., Pretti, Marques, Lima, M., Martins, B., 2021. New SARS-CoV-2 lineages could evade CD8+ T-cells response. bioRxiv 2021.03.09.434584. https://doi.org/10.1101/2021.03.09.434584

Arts, E., Brown, S., Bulir, D., Charles, T.C., Degroot, C.T., Delatolla, R., Desaulniers, J.-P., Edwards, E.A., Fuzzen, M., Gilbride, K., Gilchrist, J., Goodridge, L., Graber, T.E., Habash, M., 2022. Community Surveillance of Omicron in Ontario: Wastewater-based Epidemiology Comes of Age.https://doi.org/10.21203/RS.3.RS-1439969/V2

Carcereny, A., Martínez-Velázquez, A., Bosch, A., Allende, A., Truchado, P., Cascales, J., Romalde, J.L., Lois, M., Polo, D., Sánchez, G., Pérez-Cataluña, A., Díaz-Reolid, A., Antón, A., Gregori, J., Garcia-Cehic, D., Quer, J., Palau, M., Ruano, C.G., Pintó, R.M., Guix, S., 2021. Monitoring Emergence of the SARS-CoV-2 B.1.1.7 Variant through the Spanish National SARS-CoV-2 Wastewater Surveillance System (VATar COVID-19). Environ. Sci. Technol. 55, 11756–11766. https://doi.org/10.1021/acs.est.1c03589

Chan, C.T.-M., Leung, J.S.-L., Lee, L.-K., Lo, H.W.-H., Wong, E.Y.-K., Wong, D.S.-H., Ng, T.T.-L., Lao, H.-Y., Lu, K.K., Jim, S.H.-C., Yau, M.C.-Y., Lam, J.Y.-W., Ho, A.Y.-M., Luk, K.S., Yip, K.-T., Que, T.-L., To, K.K.-W., Siu, G.K.-H., 2022. A low-cost TaqMan minor groove binder probe-based one-step RT-qPCR assay for rapid identification of N501Y variants of SARS-CoV-2. J. Virol. Methods 299, 114333. https://doi.org/10.1016/j.jviromet.2021.114333

D’Aoust, P.M., Graber, T.E., Mercier, E., Montpetit, D., Alexandrov, I., Neault, N., Baig, A.T., Mayne, J., Zhang, X., Alain, T., Servos, M.R., Srikanthan, N., MacKenzie, M., Figeys, D., Manuel, D., Jüni, P., MacKenzie, A.E., Delatolla, R., 2021. Catching a resurgence: Increase in SARS-CoV-2 viral RNA identified in wastewater 48 h before COVID-19 clinical tests and 96 h before hospitalizations. Sci. Total Environ. 770. https://doi.org/10.1016/j.scitotenv.2021.145319

Graber, T.E., Mercier, É., Bhatnagar, K., Fuzzen, M., D’Aoust, P.M., Hoang, H.D., Tian, X., Towhid, S.T., Plaza-Diaz, J., Eid, W., Alain, T., Butler, A., Goodridge, L., Servos, M., Delatolla, R., 2021. Near real-time determination of B.1.1.7 in proportion to total SARS-CoV-2 viral load in wastewater using an allele-specific primer extension PCR strategy. Water Res. 205, 117681. https://doi.org/10.1016/J.WATRES.2021.117681

Hadfield, J., Megill, C., Bell, S.M., Huddleston, J., Potter, B., Callender, C., Sagulenko, P., Bedford, T., Neher, R.A., 2018. NextStrain: Real-time tracking of pathogen evolution. Bioinformatics 34, 4121–4123. https://doi.org/10.1093/bioinformatics/bty407

Hamouda, M., Mustafa, F., Maraqa, M., Rizvi, T., Aly Hassan, A., 2021. Wastewater surveillance for SARS-CoV-2: Lessons learnt from recent studies to define future applications. Sci. Total Environ. 759, 143493. https://doi.org/https://doi.org/10.1016/j.scitotenv.2020.143493

Johnson, B.A., Zhou, Y., Lokugamage, K.G., Vu, M.N., Bopp, N., Crocquet-Valdes, P.A., Schindewolf, C., Liu, Y., Scharton, D., Plante, J.A., Xie, X., Aguilar, P., Weaver, S.C., Shi, P.-Y., Walker, D.H., Routh, A.L., Plante, K.S., Menachery, V.D., 2021. Nucleocapsid mutations in SARS-CoV-2 augment replication and pathogenesis. bioRxiv 2021.10.14.464390. https://doi.org/10.1101/2021.10.14.464390

Katoh, K., Misawa, K., Kuma, K.I., Miyata, T., 2002. MAFFT: a novel method for rapid multiple sequence alignment based on fast Fourier transform. Nucleic Acids Res. 30, 3059–3066. https://doi.org/10.1093/NAR/GKF436

Kiryanov, S.A., Levina, T.A., Konopleva, M. V., Suslov, A.P., 2022. Identification of Hotspot Mutations in the N Gene of SARS-CoV-2 in Russian Clinical Samples That May Affect the Detection by Reverse Transcription-PCR. Diagnostics 2022, Vol. 12, Page 147 12, 147. https://doi.org/10.3390/DIAGNOSTICS12010147

Kitajima, M., Ahmed, W., Bibby, K., Carducci, A., Gerba, C.P., Hamilton, K.A., Haramoto, E., Rose, J.B., 2020. SARS-CoV-2 in wastewater: State of the knowledge and research needs. Sci. Total Environ. 739, 139076. https://doi.org/https://doi.org/10.1016/j.scitotenv.2020.139076

Kumblathan, T., Liu, Y., Uppal, G.K., Hrudey, S.E., Li, X.-F., 2021. Wastewater-Based Epidemiology for Community Monitoring of SARS-CoV-2: Progress and Challenges. ACS Environ. Au 1, 18–31. https://doi.org/10.1021/acsenvironau.1c00015

Lee, W.L., Imakaev, M., Armas, F., McElroy, K.A., Gu, X., Duvallet, C., Chandra, F., Chen, H., Leifels, M., Mendola, S., Floyd-O’Sullivan, R., Powell, M.M., Wilson, S.T., Berge, K.L.J., Lim, C.Y.J., Wu, F., Xiao, A., Moniz, K., Ghaeli, N., Matus, M., Thompson, J., Alm, E.J., 2021. Quantitative SARS-CoV-2 Alpha Variant B.1.1.7 Tracking in Wastewater by Allele-Specific RT-qPCR. Environ. Sci. Technol. Lett. 8, 675–682. https://doi.org/10.1021/acs.estlett.1c00375

Lu, X., Wang, L., Sakthivel, S.K., Whitaker, B., Murray, J., Kamili, S., Lynch, B., Malapati, L., Burke, S.A., Harcourt, J., Tamin, A., Thornburg, N.J., Villanueva, J.M., Lindstrom, S., 2020. US CDC Real-Time Reverse Transcription PCR Panel for Detection of Severe Acute Respiratory Syndrome Coronavirus 2. Emerg. Infect. Dis. 26, 1654–1665. https://doi.org/10.3201/eid2608.201246

Medema, G., Heijnen, L., Elsinga, G., Italiaander, R., Brouwer, A., 2020. Presence of SARS-Coronavirus-2 RNA in Sewage and Correlation with Reported COVID-19 Prevalence in the Early Stage of the Epidemic in The Netherlands. Environ. Sci. Technol. Lett. 7, 511–516. https://doi.org/10.1021/acs.estlett.0c00357

Pérez-Cataluña, A., Cuevas-Ferrando, E., Randazzo, W., Falcó, I., Allende, A., Sánchez, G., 2021. Comparing analytical methods to detect SARS-CoV-2 in wastewater. Sci. Total Environ. 758, 143870. https://doi.org/10.1016/J.SCITOTENV.2020.143870

Peterson, S.W., Lidder, R., Daigle, J., Wonitowy, Q., Dueck, C., Nagasawa, A., Mulvey, M.R., Mangat, C.S., 2022. RT-qPCR detection of SARS-CoV-2 mutations S 69–70 del, S N501Y and N D3L associated with variants of concern in Canadian wastewater samples. Sci. Total Environ. 810, 151283. https://doi.org/10.1016/J.SCITOTENV.2021.151283

Syed, A.M., Taha, T.Y., Tabata, T., Chen, I.P., Ciling, A., Khalid, M.M., Sreekumar, B., Chen, P.-Y., Hayashi, J.M., Soczek, K.M., Ott, M., Doudna, J.A., 2021. Rapid assessment of SARS-CoV-2 evolved variants using virus-like particles. Science eabl6184. https://doi.org/10.1126/science.abl6184

Vogels, C.B.F., Breban, M.I., Ott, I.M., Alpert, T., Petrone, M.E., Watkins, A.E., Kalinich, C.C., Earnest, R., Rothman, J.E., de Jesus, J.G., Claro, I.M., Ferreira, G.M., Crispim, M.A.E., Singh, L., Tegally, H., Anyaneji, U.J., Hodcroft, E.B., Mason, C.E., Khullar, G., Metti, J., Dudley, J.T., MacKay, M.J., Nash, M., Wang, J., Liu, C., Hui, P., Murphy, S., Neal, C., Laszlo, E., Landry, M.L., Muyombwe, A., Downing, R., Razeq, J., de Oliveira, T., Faria, N.R., Sabino, E.C., Neher, R.A., Fauver, J.R., Grubaugh, N.D., da Silva Sales, F.C., Ramundo, M.S., Candido, D.S., Silva, C.A.M., de Pinho, M.C., Coletti, T. de M., Andrade, P. dos S., de Souza, L.M., Rocha, E.C., Gomes Jardim, A.C., Manuli, E., Gaburo, N., Granato, C., Levi, J.E., Costa, S., de Souza, W.M., Salum, M.A., Pereira, R., de Souza, A., Matkin, L.E., Nogueria, M.L., Levin, A.S., Mayaud, P., Alexander, N., Souza, R., Acosta, A.L., Prete, C., Quick, J., Brady, O., Messina, J., Kraemer, M., Gouveia, N. da C., Oliva, I., de Souza, M., Lazari, C., Alencar, C.S., Thézé, J., Buss, L., Araujo, L., Cunha, M.S., Loman, N.J., Pybus, O.G., Aguiar, R.S., Wilkinson, E., Msomi, N., Iranzadeh, A., Fonseca, V., Doolabh, D., San, E.J., Mlisana, K., von Gottberg, A., Walaza, S., Allam, M., Ismail, A., Mohale, T., Glass, A.J., Engelbrecht, S., van Zyl, G., Preiser, W., Petruccione, F., Sigal, A., Hardie, D., Marais, G., Hsiao, M., Korsman, S., Davies, M.A., Tyers, L., Mudau, I., York, D., Maslo, C., Goedhals, D., Abrahams, S., Laguda-Akingba, O., Alisoltani-Dehkordi, A., Godzik, A., Wibmer, C.K., Sewell, B.T., Lourenço, J., Kosakovsky Pond, S.L., Weaver, S., Giovanetti, M., Alcantara, L.C.J., Martin, D., Bhiman, J.N., Williamson, C., 2021. Multiplex qPCR discriminates variants of concern to enhance global surveillance of SARS-CoV-2. PLoS Biol. 19, e3001236. https://doi.org/10.1371/journal.pbio.3001236

Wu, F., Zhang, J., Xiao, A., Gu, X., Lee, W.L., Armas, F., Kauffman, K., Hanage, W., Matus, M., Ghaeli, N., Endo, N., Duvallet, C., Poyet, M., Moniz, K., Washburne, A.D., Erickson, T.B., Chai, P.R., Thompson, J., Alm, E.J., 2020. SARS-CoV-2 Titers in Wastewater Are Higher than Expected from Clinically Confirmed Cases. mSystems 5. https://doi.org/10.1128/msystems.00614-20

Wu, H., Xing, N., Meng, K., Fu, B., Xue, W., Dong, P., Tang, W., Xiao, Y., Liu, G., Luo, H., Zhu, W., Lin, X., Meng, G., Zhu, Z., 2021. Nucleocapsid mutations R203K/G204R increase the infectivity, fitness, and virulence of SARS-CoV-2. Cell Host Microbe 29, 1788-1801.e6. https://doi.org/10.1016/j.chom.2021.11.005

Wurtzer, S., Waldman, P., Levert, M., Mouchel, J.M., Gorgé, O., Boni, M., Maday, Y., consortium, O., Marechal, V., Moulin, L., 2021. Monitoring the propagation of SARS CoV2 variants by tracking identified mutation in wastewater using specific RT-qPCR. medRxiv 2021.03.10.21253291. https://doi.org/10.1101/2021.03.10.21253291

Yaniv, K., Ozer, E., Shagan, M., Lakkakula, S., Plotkin, N., Bhandarkar, N.S., Kushmaro, A., 2021. Direct RT-qPCR assay for SARS-CoV-2 variants of concern (Alpha, B.1.1.7 and Beta, B.1.351) detection and quantification in wastewater. Environ. Res. 201, 111653. https://doi.org/10.1016/j.envres.2021.111653

Ye, J., Coulouris, G., Zaretskaya, I., Cutcutache, I., Rozen, S., Madden, T.L., 2012. Primer-BLAST: a tool to design target-specific primers for polymerase chain reaction. BMC Bioinformatics 13, 134. https://doi.org/10.1186/1471-2105-13-134/FIGURES/5

